# Chikungunya Seroprevalence among Patients Presenting with Febrile illnesses in selected health facilities in Mt. Elgon region, Kenya

**DOI:** 10.1101/2024.04.26.24306414

**Authors:** Sheila Kageha, Joyce M. Ngoi, Toru Kubo, Kouichi Morita, Matilu Mwau

**Affiliations:** Kenya Medical Research Institute, Kenya; West African Centre for Cell Biology of Infectious Pathogens (WACCBIP), College of Basic and Applied Sciences, University of Ghana, Accra, Ghana; Department of Virology, Institute of Tropical Medicine, Nagasaki University, Japan

**Author notes:** Corresponding Author: Sheila Kageha, Mail, P.O. Box 54628 Nairobi-00200, Kenya.

**Keywords:** Chikungunya virus (CHIKV), Seroprevalence survey, Kenya, Mt. Elgon

## Abstract

**Background:** Chikungunya is an emerging epidemic-prone vector-borne disease of considerable significance globally. Infection with chikungunya virus induces an acute illness characterized by fever and painful arthralgia, which can evolve to chronic arthritis and rheumatism especially in elderly patients. Whereas febrile illness and arthralgia are common clinical presentations amongst residents of Mt. Elgon, the role of chikungunya virus as a causative agent is undocumented. This study was carried out to determine the prevalence of IgA, IgM and IgG antibodies against Chikungunya Virus (CHIKV) antigens in patients presenting with acute febrile illnesses in Mt. Elgon region, Kenya.

**Methods:** This was a cross-sectional seroprevalence study on febrile patients visiting Endebes, Andersen and Kitale County Referal Hospitals. Sociodemographic data was collected whenever possible. Serum samples were collected and screened using Indirect ELISA for 1gG+IgM+IgA antibodies. Sera that tested positive by ELISA were subjected to standard plaque reduction neutralization assays (PRNT) performed on monolayer cultures of Vero E6 cells for confirmation.

**Results:** By ELISA, a total of 317/1359 (23.33%) sera were positive for CHIKV antibodies. Of the 317 positive sera, 305 (96.21%) were of sufficient quantity and were subjected to PRNT. Ultimately, 127 (9.3%) samples tested positive for CHIKV neutralising antibodies by PRNT.

**Conclusion:** These findings suggest active circulation of CHIKV in Mt. Elgon, even though it has previously been considered a non-endemic region for the virus. There is need to closely monitor and continuously put in place surveillance strategies to prevent probable potential outbreaks in the future.

## Introduction

Chikungunya virus (CHIKV), an alphavirus of the Togaviridae family [1-4], was originally isolated in 1952-53 from the serum of a febrile patient during an outbreak that occurred in the Makonde Plateau in the southern region of Tanzania [5]. It has since spread, infecting millions of people globally. CHIKV infection is commonly characterised by acute fever, headaches and painful arthralgia, which can evolve to chronic arthritis and rheumatism especially in elderly patients[5-7]. These symptoms are often clinically indistinguishable from those caused by other arboviruses such as dengue, Onyong’ nyong’ (ONNV) and West Nile viruses (WNV). Similarities in clinical symptoms often lead to missed opportunities to detect, and consequent underreporting of CHIKV infection in areas endemic to arbovirus infections [8, 9]. CHIKV has become a global public health threat with reported cases of neurologic involvement, fulminant hepatitis, and neonatal encephalopathy[10, 11]. Neurological forms of the disease and severe cases have also been reported in the Réunion island outbreak of 2005-2006 and in Italy [12-14].

Currently there are no licensed specific antiviral drugs or vaccines to prevent CHIKV infection. Treatment is mainly supportive and entails the use of analgesic drugs in combination with non-steroidal anti-inflammatory drugs for symptomatic relief [15-17]. Patients infected with CHIKV develop robust innate and adaptive immune responses that help in viral clearance and protection. Neutralising anti-CHIKV IgG persists for at least 21 months [18-20].

CHIKV circulates in a sylvatic cycle between non-human primates/mammalian reservoir hosts and *Aedes spp*. of mosquitoes [21]. *Aedes aegypti* is responsible for CHIKV endemicity in the tropical and sub-tropical regions while *Aedes albopictus* has been associated with spread in temperate regions including southern Europe, the Caribbean and southern and eastern regions of the USA. The virus is transmitted to human hosts via infectious bites from *Aedes spp*. of mosquitoes [22, 23]. There has been increased frequency of CHIKV outbreaks in previously non-endemic regions, which has been attributed to the expansion of mosquito vector species and genetic adaptation of the virus that enhances its transmissibility [24-26].

CHIKV originally endemic in Africa is known for its wide geographic distribution. In Africa, there have been reports of human infection from Angola, Democratic Republic of Congo, Mozambique, Gabon, Nigeria, Southern Africa, Tanzania, Uganda, and Burundi [27-31]. Chikungunya has since spread to Asia including India and in the island countries of the Indian Ocean, Europe and America. In America, the virus has been identified in 45 states, and more than 2.9 million suspected and confirmed cases and 296 deaths have been reported [32]. In Asia, CHIKV was first isolated in Bangkok, Thailand in 1958 [29, 33].

The disease was formerly thought to be self-limiting until the Reunion Island outbreaks in 2005-2006 that witnessed several fatalities. Brazil has also experienced fatalities resulting from Chikungunya outbreak [34].

In Kenya, the first outbreaks of CHIKV infection were recorded in Lamu and Mombasa in 2004[35, 36]. Indeed, genetic analysis of the virus responsible for the Indian Ocean epidemic showed that it originated in the Kenyan Coast. CHIKV vectors are commonly found in diverse habitats, and there is reason to believe that CHIKV outbreaks have occurred before. Mt. Elgon region in Trans Nzoia County has substantial populations of reservoir hosts, high vector infestation and favourable climatic conditions for transmission of arboviruses. Furthermore, encroachment of human populations into forested areas for agricultural land has led to increased exposure to sylvatic mosquitoes that vector these viruses. This study aimed to determine the seroprevalence of CHIKV infection in febrile patient population presenting in selected health facilities in the region.

## Materials and Methods

### Study sites and specimen collection

This was a cross-sectional study conducted to determine the seroprevalence of CHIKV in febrile patients visiting health facilities in Mt. Elgon region. Mt. Elgon ecosystem comprises a forest reserve and a national park. The surrounding areas are heavily forested. The slopes of the mountain nearby have several caves, infested with bats and other rodents. The livelihoods of the population is mainly linked to agricultural production. This has seen encroachment of the forest for cultivation and exploitation of forest products such as fuelwood, medicinal herbs, posts and poles, grazing and hunting for wild game.

The study was carried out in three health facilities located in the region: Andersen Medical centre (AMC), Endebes District hospital (END) and Kitale County Referral hospital (KCRH within the County. These facilities serve a large part of Trans Nzoia County. With consent, venous blood samples were collected from patients ≥ 5 years of age presenting with acute febrile illnesses. Structured questionnaires were used to collect socio-demographic and clinical information from a subset of study participants. Ethical approval for the study protocol was sought from Kenya Medical Research Institute Ethical Review Committee under SSC No.1698. All samples were anonymized at collection by assigning unique codes.

Storage was at −20° C at the facilities. Samples were transported in dry ice for processing at KEMRI, where sera were separated, aliquoted into vials and stored at −80°C.

### Laboratory Assays

Sera were tested for 1gG+IgM+IgA antibodies against CHIKV and neutralizing antibodies against CHIKV for those that tested positive by ELISA. All sera were first tested for antibodies against CHIKV antigens by enzyme-linked immunosorbent assay (ELISA). An in-house indirect ELISA was performed according to the methods described in the Igarashi Technical Manual (2000) with some modifications. Purified CHIK viruses from infected culture fluid of C6/36 cells were used as antigens. Polyethylene glycol and NaCl was used to concentrate the virus antigens, which were then purified using sucrose-gradient ultracentrifugation at 50000g, for 14h at 4 °C (Bundo and Igarashi, 1983) [50]. The viral antigens were diluted at 250ng/100 μL with phosphate buffered saline (PBS). High-protein-binding 96-well microplates (Maxisorp; Nulgenunc international, Roskilde, Denmark) were coated with 250ng/100 μL/well viral antigen, wrapped and incubated overnight at 4 °C. Blocking was done by adding 100μL of PBS, 3% FCS (PBS-F) to all the wells and then incubated for 30 min at RT. The test sera were diluted at 1/1000 with PBS-F and added in duplicates to the pre-coated plates and incubation done at 37°C for 1 hr.

Control sera (both positive and negative controls) were run on each plate. After incubation, the plate was washed three times with PBS, 0.05% Tween-20. 100 μL/well of 1:5000 diluted horseradish-conjugated goat anti-human IgG+M+A (American Qualex, A139PN) [37] was added to the wells and incubated at 37 °C for 1h followed by washing three times. A substrate solution consisting of 5mg O-Phenylenediamine dihydrochloride (OPD) (Sigma Chemical, St. Louis, MO) was added and incubated for 15 min at RT in the dark. 100 μL/well of 5N Sulfuric acid was then added to stop further reaction. Absorbance (OD) of each well was determined using spectrophotometer at 492nm. OD specific for CHKV was calculated as follows [(Mean OD of virus coated wells) – (Mean OD of PBS-F coated wells)]. A serum sample was deemed positive if the mean OD difference was ≥1.

### Virus neutralization assay

CHIKV antigens obtained from bulk virus cultures and purified by sucrose density gradient ultracentrifugation were used in this assay. Titres of virus stocks were determined by plaque assays in Vero cells expressed as plaque---forming units (PFU) per mL. All patient sera positive by IgG+IgM+IgA ELISA were subsequently subjected to PRNT (protocol adapted from Igarashi Technical Manual, 2000). This assay was used to determine the presence of CHIKV specific neutralizing antibodies in the test sera. The PRNTs were performed using Vero E6 cells. CHIKV prototype strain (S 27) was used for this experiment. PRNT plates were prepared by seeding Vero cells at a concentration of 1 × 10^5^ cells/ml into 6-well plates at a volume of 3 ml/well. The cells were cultured in Growth Medium (EMEM (Gibco), 10% FCS (Gibco), L-glutamine P/S (Gibco), 0.2mM NEAA (Gibco), NaHCO3 supplemented for 1 day at 37ºC, 5% CO_2_ until the cells had attained 80% confluence. Test serum was diluted with Maintenance Medium (MM) (EMEM, 2% FCS, L-glutamine, P/S, NEAA, NaHCO3 supplemented) into 1:10 dilutions. At the same time, standard virus dilution of 1000 PFU/ml with MM was prepared. An equal volume of serum dilution (100μL) and standard virus dilution (100μL) was mixed. For positive control wells, equal volumes of MM and standard virus dilution were mixed. The virus-serum mixture was then incubated for 1 hour at 37ºC. After 1h incubation of mixture, 2.5 ml of culture medium from each well of 6-well plates was aspirated and 100 μl/well of serum/virus mixture added to Vero cells in duplicate wells. The plates were then incubated for 1.5 - 2 hours in the 37ºC, 5% CO_2_ incubator. A total of 3ml of overlay medium (EMEM, 2% FCS, 1.4% Methylcellulose, P/S supplemented) was added into each well, and incubated at 37ºC, 5% CO_2_ for 2-3 days with daily observation. Harvesting involved discarding of overlay medium and fixing plates by adding 1 ml of 10% formalin in PBS over the cell and incubation for 1 hour at room temperature (rt) in the safety cabinet under UV light. After incubation, the plates were then washed gently with tap water at least twice, absorbed on paper towel and staining solution (0.5 ml of 1% Crystal Violet solution in water) added. The stain was discarded after 5-10 minutes at room temperature and plates washed gently with tap water. The plates were air-dried at room temperature and plaques counted for each set of duplicate wells. The percentage reduction was calculated by comparing with the positive control virus well (100% plaque formation). Sera were initially tested at a dilution of 1:20 and those that reduced the number of plaques by ≥75% (PRNT_70_) were titrated. More than 90% plaque reduction (PRNT_90_) was regarded as positive and titers were expressed as the reciprocal of serum dilutions yielding ≥90% reduction in the number of plaques.

### Data analysis

Data analyses were performed using STATA Version 14 software (Stata Corp LP, College Station, Texas, United States). Chi-square test was performed for potential association. Two-sided p values were reported, and statistical significance was taken as p < 0.05.

## Results

A total of 1,398 samples were collected from febrile patients in the selected health facilities and screened for CHIKV antibodies. Thirty-nine (39, 2.79%) patient sera were excluded from analysis for various reasons including insufficient sera volume and compromised sample integrity.

The main inclusion criterion for this study was fever. The minimum temperature was 38°C while the maximum was 40.5°C with a mean of 39.1°C. The median duration of fever was nine days. Forty-three patients complained of muscle pains. Other complaints included: jaundice, body pains, abdominal pains, diarrhoea and bleeding.

Demographic and clinical data was available for 782/1,359 (57.54%) samples: 507 (37.31%) were collected from females and 275 (20.24%) from males (Table 1). The average age of the participants was 31.6 years (Median 28, range 5-91 years). There was no significant difference in ages between males and females, (median 31.4 and 31.7 respectively, *p*>0.6). Data on residence of patients was available for 777 samples. Majority of the people resided in formerly Rift Valley Province (83.71%), followed by Western Province (2.1%), Eastern Uganda (11.2%) while the rest (3%) were non-residents. Of the 1,359 samples processed successfully, 689 (50.70%) were from KCRH, 463 (34.07%) from AMC and 207 (15.23%) from END. A total of 317 (23.34%) samples tested antibody positive by ELISA.

**Table 1:**
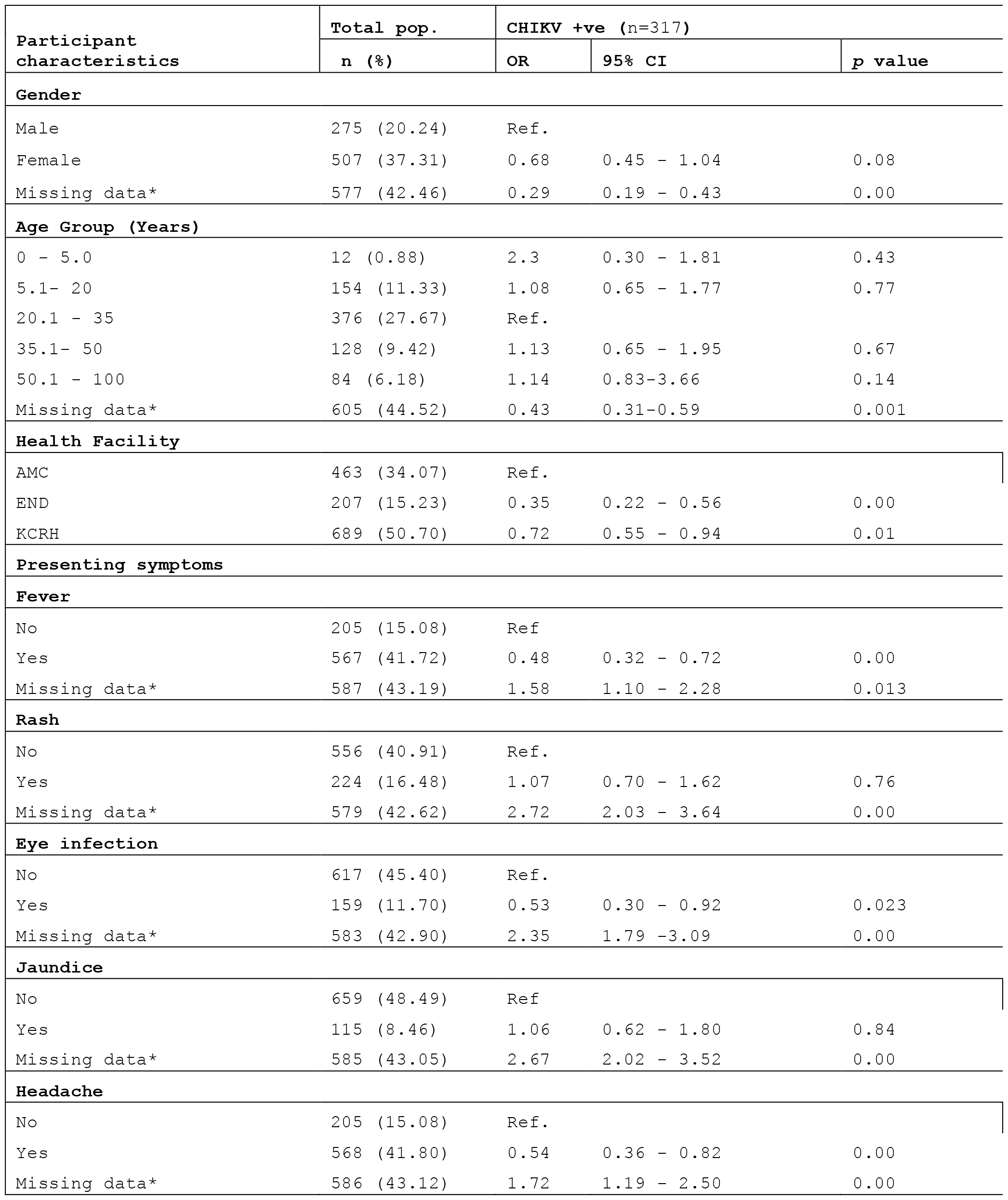
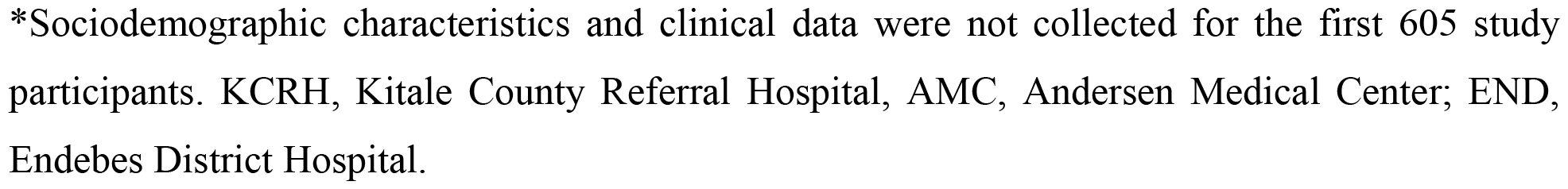
Poisson Regression Analysis for Chikungunya Virus (CHIKV) Seropositivity among febrile patients visiting selected health facilities in Mt. Elgon Region.

| Participant characteristics | Total pop. | CHIKV +ve (n=317) |  |  |
| --- | --- | --- | --- | --- |
|  | n (%) | OR | 95% CI | p value |
| **Gender** |  |  |  |  |
| Male | 275 (20.24) | Ref. |  |  |
| Female | 507 (37.31) | 0.68 | 0.45 - 1.04 | 0.08 |
| Missing data* | 577 (42.46) | 0.29 | 0.19 - 0.43 | 0.00 |
| **Age Group (Years)** |  |  |  |  |
| 0 - 5.0 | 12 (0.88) | 2.3 | 0.30 - 1.81 | 0.43 |
| 5.1- 20 | 154 (11.33) | 1.08 | 0.65 - 1.77 | 0.77 |
| 20.1 - 35 | 376 (27.67) | Ref. |  |  |
| 35.1- 50 | 128 (9.42) | 1.13 | 0.65 - 1.95 | 0.67 |
| 50.1 - 100 | 84 (6.18) | 1.14 | 0.83-3.66 | 0.14 |
| Missing data* | 605 (44.52) | 0.43 | 0.31-0.59 | 0.001 |
| **Health Facility** |  |  |  |  |
| AMC | 463 (34.07) | Ref. |  |  |
| END | 207 (15.23) | 0.35 | 0.22 - 0.56 | 0.00 |
| KCRH | 689 (50.70) | 0.72 | 0.55 - 0.94 | 0.01 |
| **Presenting symptoms** |  |  |  |  |
| **Fever** |  |  |  |  |
| No | 205 (15.08) | Ref |  |  |
| Yes | 567 (41.72) | 0.48 | 0.32 - 0.72 | 0.00 |
| Missing data* | 587 (43.19) | 1.58 | 1.10 - 2.28 | 0.013 |
| **Rash** |  |  |  |  |
| No | 556 (40.91) | Ref. |  |  |
| Yes | 224 (16.48) | 1.07 | 0.70 - 1.62 | 0.76 |
| Missing data* | 579 (42.62) | 2.72 | 2.03 - 3.64 | 0.00 |
| **Eye infection** |  |  |  |  |
| No | 617 (45.40) | Ref. |  |  |
| Yes | 159 (11.70) | 0.53 | 0.30 - 0.92 | 0.023 |
| Missing data* | 583 (42.90) | 2.35 | 1.79 -3.09 | 0.00 |
| **Jaundice** |  |  |  |  |
| No | 659 (48.49) | Ref |  |  |
| Yes | 115 (8.46) | 1.06 | 0.62 - 1.80 | 0.84 |
| Missing data* | 585 (43.05) | 2.67 | 2.02 - 3.52 | 0.00 |
| **Headache** |  |  |  |  |
| No | 205 (15.08) | Ref. |  |  |
| Yes | 568 (41.80) | 0.54 | 0.36 - 0.82 | 0.00 |
| Missing data* | 586 (43.12) | 1.72 | 1.19 - 2.50 | 0.00 |

\*Sociodemographic characteristics and clinical data were not collected for the first 605 study participants. KCRH, Kitale County Referral Hospital, AMC, Andersen Medical Center; END, Endebes District Hospital.

The odds of exposure were determined for gender, health facilities, age of participants and presenting symptoms (Table 1). The odds of being seropositive for CHIKV were significantly lower in END and KCRH when compared with AMC, in those with fever when compared to those without, those with eye infection and headache when compared to those without (p<0.05).

Of the 317 samples that tested antibody positive by ELISA, 305 (96.21%) were further subjected to PRNT for CHIKV-specific neutralizing antibodies. A total of 123 (40.33%) exhibited neutralizing activities for CHIKV antigens. An additional four (4) samples that were borderline negative by ELISA tested PRNT positive (Table 2).

**Table 2:** ELISA versus PRNT Results for CHIKV.

| ELISA Status | CHIKV PRNT |  |  |
| --- | --- | --- | --- |
|  | Neutralizing | None Neutralizing | Total |
| NEG | 4<br>80.00 | 1<br>20.00 | 5<br>100.00 |
| POS | 123<br>41.00 | 177<br>59.00 | 300<br>100.00 |
| Total | 127<br>41.64 | 178<br>58.36 | 305<br>100.00 |

## Discussion

Our study observed an overall seroprevalence of 23.34% (317) by ELISA. PRNT results were positive for 9.35% (127) of all samples, confirming CHIKV activity in the region. Mt. Elgon region exhibits extraordinary biodiversity due to the presence of forests, rivers, high levels of humidity and vertebrate reservoirs that provides potential *Aedes spp*. breeding habitats. These ecological settings provide opportunities for multiplication and the spread of vector-borne viruses in the region.

Agriculture is the main economic activity in the region. This has seen encroachment on forest reserves for cultivation and exploitation of forest products such as fuelwood, medicinal herbs, timber, grazing and hunting for wild game; increasing exposure to the sylvatic mosquitoes that vector these viruses. Aedes spp. responsible for CHIKV transmission have been reported in Western Kenya in previous studies [38, 39].

The high seropositivity in this study compares with findings from past studies in the area, indicating that alphaviruses, though subtle, are actively circulating in the region [40]. Other studies in Cameroon, Benin, Mozambique and Guinea have shown similar results, which suggest a significant and unrecognized CHIKV circulation in Africa[41-44].

In this study, we could not ascertain with confidence whether the seropositivity observed was due to present or convalescent CHIKV and/or ONNV infections. Little is known about the lifeline of circulatory CHIKV specific antibodies, beyond what is generally known about the kinetics of IgG and IgM antibodies with other immune correlates. However, herd immunity has been demonstrated in populations with constant contact with vectors, this could explain the observed neutralization [44, 45].

Of interest, were samples that tested positive by ELISA but failed to neutralize the CHIKV antigens by PRNT 178/305 (58.36%). This suggests possible IgM/IgG cross-reactivity to closely related alphaviruses circulating in the region. CHIKV belongs within the Semliki Forest virus (SFV) antigenic serocomplex and therefore exhibits serological cross-reactivity[45, 46]. Co-infections with more than one etiological agent have been shown to complicate diagnosis, severity and management of these viruses in patients [47, 48]. More diverse studies to assess CHKV antibody persistence and the likelihood of cross-protection against CHIKV infection due to previous alphavirus infection could provide better insights. Findings from related studies have indicated multiple genotypes that could have effects on serological outcomes and epidemic potential[27, 49].

In Kenya, especially in malaria endemic regions, malaria is frequently diagnosed as the cause of acute undifferentiated febrile illnesses while other causes like arboviruses are often overlooked. This study suggests that CHIKV is an important cause of fever in Mt. Elgon and should be considered as a differential diagnosis in clinical practise.

This study had limitations such as incomplete and insufficient demographics and clinical information that hampered analysis for odds of exposure among the population.

## Conclusion

From this study, we conclude that CHIKV commonly infects residents of Mt. Elgon, and is under-recognized as a cause of morbidity in this population. Therefore, periodic surveillance is necessary to assess the viral burden in the population and devise effective public health interventions to tackle the challenge. Vector control measures are also needed to curb these infections.

## Data Availability

All the data is in the manuscript. However, any extra data, if required can be made available on request.

## Acknowledgement

This study was supported by Nagasaki University of Infectious and Tropical Medicine in collaboration with the Kenya Medical Research Institute. This study appreciates the Deputy Director, Centre for Virus research and the technical staff for support.

## References

1. Higashi N, Matsumoto A, Tabata K, Nagatomo Y: Electron microscope study of development of Chikungunya virus in green monkey kidney stable (VERO) cells. Virology 1967, 33(1):69-p55.

2. Powers AM, Brault AC, Shirako Y, Strauss EG, Kang W, Strauss JH, Weaver SC: Evolutionary relationships and systematics of the alphaviruses. Journal of virology 2001, 75(21):10118– 10131.

3. Strauss JH, Strauss EG: The alphaviruses: gene expression, replication, and evolution. Microbiological reviews 1994, 58(3):491–562.

4. Lahariya C, Pradhan SK: Emergence of chikungunya virus in Indian subcontinent after 32 years: A review. Journal of vector borne diseases 2006, 43(4):151–160.

5. Robinson MC: An epidemic of virus disease in Southern Province, Tanganyika Territory, in 1952-53. I. Clinical features. Transactions of the Royal Society of Tropical Medicine and Hygiene 1955, 49(1):28–32.

6. Miner JJ, Aw-Yeang HX, Fox JM, Taffner S, Malkova ON, Oh ST, Kim AHJ, Diamond MS, Lenschow DJ, Yokoyama WM: Chikungunya viral arthritis in the United States: a mimic of seronegative rheumatoid arthritis. Arthritis & rheumatology (Hoboken, NJ) 2015, 67(5):1214–1220.

7. Weaver SC: Arrival of chikungunya virus in the new world: prospects for spread and impact on public health. PLoS neglected tropical diseases 2014, 8(6):e2921.

8. Carey DE: Chikungunya and dengue: a case of mistaken identity? Journal of the history of medicine and allied sciences 1971, 26(3):243–262.

9. Seneviratne SL, Perera J: Fever epidemic moves into Sri Lanka. BMJ (Clinical research ed) 2006, 333(7580):1220–1221.

10. Rampal, Sharda M, Meena H: Neurological complications in Chikungunya fever. The Journal of the Association of Physicians of India 2007, 55:765–769.

11. Wielanek AC, Monredon JD, Amrani ME, Roger JC, Serveaux JP: Guillain-Barré syndrome complicating a Chikungunya virus infection. Neurology 2007, 69(22):2105–2107.

12. Tournebize P, Charlin C, Lagrange M: [Neurological manifestations in Chikungunya: about 23 cases collected in Reunion Island]. Revue neurologique 2009, 165(1):48–51.

13. Lemant J, Boisson V, Winer A, Thibault L, André H, Tixier F, Lemercier M, Antok E, Cresta MP, Grivard P et al: Serious acute chikungunya virus infection requiring intensive care during the Reunion Island outbreak in 2005-2006. Critical care medicine 2008, 36(9):2536–2541.

14. Elsinga J, Gerstenbluth I, van der Ploeg S, Halabi Y, Lourents NT, Burgerhof JG, van der Veen HT, Bailey A, Grobusch MP, Tami A: Long-term Chikungunya Sequelae in Curaçao: Burden, Determinants, and a Novel Classification Tool. The Journal of infectious diseases 2017, 216(5):573–581.

15. Brighton SW: Chloroquine phosphate treatment of chronic Chikungunya arthritis. An open pilot study. South African medical journal = Suid-Afrikaanse tydskrif vir geneeskunde 1984, 66(6):217–218.

16. Staikowsky F, Le Roux K, Schuffenecker I, Laurent P, Grivard P, Develay A, Michault A: Retrospective survey of Chikungunya disease in Reunion Island hospital staff. Epidemiol Infect 2008, 136(2):196–206.

17. De Lamballerie X, Boisson V, Reynier JC, Enault S, Charrel RN, Flahault A, Roques P, Le Grand R: On chikungunya acute infection and chloroquine treatment. Vector Borne Zoonotic Dis 2008, 8(6):837–839.

18. Kam YW, Lee WW, Simarmata D, Harjanto S, Teng TS, Tolou H, Chow A, Lin RT, Leo YS, Rénia L et al: Longitudinal analysis of the human antibody response to Chikungunya virus infection: implications for serodiagnosis and vaccine development. Journal of virology 2012, 86(23):13005–13015.

19. Seymour RL, Adams AP, Leal G, Alcorn MD, Weaver SC: A Rodent Model of Chikungunya Virus Infection in RAG1 −/− Mice, with Features of Persistence, for Vaccine Safety Evaluation. PLoS neglected tropical diseases 2015, 9(6):e0003800.

20. Smith SA, Silva LA, Fox JM, Flyak AI, Kose N, Sapparapu G, Khomandiak S, Ashbrook AW, Kahle KM, Fong RH et al: Isolation and Characterization of Broad and Ultrapotent Human Monoclonal Antibodies with Therapeutic Activity against Chikungunya Virus. Cell host & microbe 2015, 18(1):86–95.

21. Pialoux G, Gaüzère BA, Jauréguiberry S, Strobel M: Chikungunya, an epidemic arbovirosis. The Lancet Infectious diseases 2007, 7(5):319–327.

22. Diallo M, Thonnon J, Traore-Lamizana M, Fontenille D: Vectors of Chikungunya virus in Senegal: current data and transmission cycles. The American journal of tropical medicine and hygiene 1999, 60(2):281–286.

23. Arias-Goeta C, Mousson L, Rougeon F, Failloux AB: Dissemination and transmission of the E1-226V variant of chikungunya virus in Aedes albopictus are controlled at the midgut barrier level. PloS one 2013, 8(2):e57548.

24. Tsetsarkin KA, Vanlandingham DL, McGee CE, Higgs S: A single mutation in chikungunya virus affects vector specificity and epidemic potential. PLoS pathogens 2007, 3(12):e201.

25. Horwood PF, Buchy P: Chikungunya. Revue scientifique et technique (International Office of Epizootics) 2015, 34(2):479–489.

26. Tsetsarkin KA, Weaver SC: Sequential adaptive mutations enhance efficient vector switching by Chikungunya virus and its epidemic emergence. PLoS pathogens 2011, 7(12):e1002412.

27. Burt FJ, Chen W, Miner JJ, Lenschow DJ, Merits A, Schnettler E, Kohl A, Rudd PA, Taylor A, Herrero LJ et al: Chikungunya virus: an update on the biology and pathogenesis of this emerging pathogen. The Lancet Infectious diseases 2017, 17(4):e107–e117.

28. Sergon K, Njuguna C, Kalani R, Ofula V, Onyango C, Konongoi LS, Bedno S, Burke H, Dumilla AM, Konde J et al: Seroprevalence of Chikungunya virus (CHIKV) infection on Lamu Island, Kenya, October 2004. The American journal of tropical medicine and hygiene 2008, 78(2):333–337.

29. Bhatia R, Narain JP: Re-emerging chikungunya fever: some lessons from Asia. Tropical medicine & international health : TM & IH 2009, 14(8):940–946.

30. Pastorino B, Muyembe-Tamfum JJ, Bessaud M, Tock F, Tolou H, Durand JP, Peyrefitte CN: Epidemic resurgence of Chikungunya virus in democratic Republic of the Congo: identification of a new central African strain. Journal of medical virology 2004, 74(2):277–282.

31. D’Ortenzio E, Grandadam M, Balleydier E, Jaffar-Bandjee MC, Michault A, Brottet E, Baville M, Filleul L: A226V strains of Chikungunya virus, Réunion Island, 2010. Emerging infectious diseases 2011, 17(2):309–311.

32. Centers for Disease Control and Prevention tUSoACvgdAfhwcgcgihLu.

33. Arankalle VA, Shrivastava S, Cherian S, Gunjikar RS, Walimbe AM, Jadhav SM, Sudeep AB, Mishra AC: Genetic divergence of Chikungunya viruses in India (1963-2006) with special reference to the 2005-2006 explosive epidemic. The Journal of general virology 2007, 88(Pt 7):1967–1976.

34. Brito CAA: Alert: Severe cases and deaths associated with Chikungunya in Brazil. Revista da Sociedade Brasileira de Medicina Tropical 2017, 50(5):585–589.

35. Kariuki Njenga M, Nderitu L, Ledermann JP, Ndirangu A, Logue CH, Kelly CHL, Sang R, Sergon K, Breiman R, Powers AM: Tracking epidemic Chikungunya virus into the Indian Ocean from East Africa. The Journal of general virology 2008, 89(Pt 11):2754–2760.

36. Sang RC, Ahmed O, Faye O, Kelly CL, Yahaya AA, Mmadi I, Toilibou A, Sergon K, Brown J, Agata N et al: Entomologic investigations of a chikungunya virus epidemic in the Union of the Comoros, 2005. The American journal of tropical medicine and hygiene 2008, 78(1):77–82.

37. Peyrefitte CN, Rousset D, Pastorino BA, Pouillot R, Bessaud M, Tock F, Mansaray H, Merle OL, Pascual AM, Paupy C et al: Chikungunya virus, Cameroon, 2006. Emerg Infect Dis 2007, 13(5):768–771.

38. Eastwood G, Sang RC, Guerbois M, Taracha ELN, Weaver SC: Enzootic Circulation of Chikungunya Virus in East Africa: Serological Evidence in Non-human Kenyan Primates. The American journal of tropical medicine and hygiene 2017, 97(5):1399–1404.

39. Ajamma YU, Onchuru TO, Ouso DO, Omondi D, Masiga DK, Villinger J: Vertical transmission of naturally occurring Bunyamwera and insect-specific flavivirus infections in mosquitoes from islands and mainland shores of Lakes Victoria and Baringo in Kenya. 2018, 12(11):e0006949.

40. Mease LE, Coldren RL, Musila LA, Prosser T, Ogolla F, Ofula VO, Schoepp RJ, Rossi CA, Adungo N: Seroprevalence and distribution of arboviral infections among rural Kenyan adults: a cross-sectional study. Virology journal 2011, 8:371.

41. Adam A, Seidahmed OM, Weber C, Schnierle B, Schmidt-Chanasit J, Reiche S, Jassoy C: Low Seroprevalence Indicates Vulnerability of Eastern and Central Sudan to Infection with Chikungunya Virus. Vector borne and zoonotic diseases (Larchmont, NY) 2016, 16(4):290–291.

42. António VS, Muianga AF, Wieseler J, Pereira SA, Monteiro VO, Mula F, Chelene I, Chongo IS, Oludele JO, Kümmerer BM et al: Seroepidemiology of Chikungunya Virus Among Febrile Patients in Eight Health Facilities in Central and Northern Mozambique, 2015-2016. Vector borne and zoonotic diseases (Larchmont, NY) 2018, 18(6):311–316.

43. Jentes ES, Robinson J, Johnson BW, Conde I, Sakouvougui Y, Iverson J, Beecher S, Bah MA, Diakite F, Coulibaly M et al: Acute arboviral infections in Guinea, West Africa, 2006. The American journal of tropical medicine and hygiene 2010, 83(2):388–394.

44. Antonio VS, Amade NA, Muianga AF, Ali S, Monteiro V, Mula F, Chelene I, Oludele J: Retrospective investigation of antibodies against chikungunya virus (CHIKV) in serum from febrile patients in Mozambique, 2009-2015: Implications for its prevention and control. 2019, 14(3):e0213941.

45. LaBeaud AD, Banda T, Brichard J, Muchiri EM, Mungai PL, Mutuku FM, Borland E, Gildengorin G, Pfeil S, Teng CY et al: High rates of o’nyong nyong and Chikungunya virus transmission in coastal Kenya. PLoS neglected tropical diseases 2015, 9(2):e0003436.

46. Powers AM, Logue CH: Changing patterns of chikungunya virus: re-emergence of a zoonotic arbovirus. The Journal of general virology 2007, 88(Pt 9):2363–2377.

47. Yong LS, Koh KC: A case of mixed infections in a patient presenting with acute febrile illness in the tropics. Case reports in infectious diseases 2013, 2013:562175.

48. Makiala-Mandanda S, Ahuka-Mundeke S, Abbate JL, Pukuta-Simbu E, Nsio-Mbeta J, Berthet N, Leroy EM, Becquart P, Muyembe-Tamfum JJ: Identification of Dengue and Chikungunya Cases Among Suspected Cases of Yellow Fever in the Democratic Republic of the Congo. Vector borne and zoonotic diseases (Larchmont, NY) 2018, 18(7):364–370.

49. Zeller H, Van Bortel W, Sudre B: Chikungunya: Its History in Africa and Asia and Its Spread to New Regions in 2013-2014. The Journal of infectious diseases 2016, 214(suppl 5):S436–s440.

